# Evaluation of Elicit as an adjunct search, screening, and data extraction tool for systematic reviews

**DOI:** 10.64898/2026.10.02.26364621

**Authors:** Isabel Wei, Jiya N Shah, Anisha Miah, Colin Hawco, Fiona Coutts, Laura Sikstrom, Stephanie H Ameis, Aristotle N Voineskos, Tristan Glatard, Erin W Dickie

## Abstract

**Background:** Systematic reviews are critical for informing evidence-based medicine, policy, and future research. However, conducting a systematic review is labour- and cost-intensive and error-prone. Elicit is an AI tool that claims to replicate the systematic review process.

**Methods:** Our study investigated Elicit’s ability to replicate a systematic review by comparing its results with those of an existing systematic review recently published by our research group. We calculated accuracy, precision, sensitivity, error rate, and person-time savings at each stage of the systematic review.

**Results:** At search, Elicit had poor overlap, finding 158 of the 5285 studies. For abstract and full-text screening, Elicit had moderate sensitivity and accuracy but poor precision, with a skew toward false positives. Elicit’s data extraction tool was promising, with sentence-specific citations, and provided helpful information for 85% of values even when it did not find exact matches. Elicit offered person-time savings, taking 3.16 hours to complete the full systematic review process.

**Conclusions:** Elicit shows promise as an adjunct screening and extraction AI tool, but in its current form, it is not ready as a standalone tool to replace the systematic review process.

## Introduction

A systematic review is the synthesis and critical appraisal of evidence from primary research studies that answer a specifically formulated research question [1]. They are integral to the implementation of clinical medicine through compiling and evaluating existing research [2]. After developing a search strategy and screening protocol, the process involves four main steps: title and abstract screening, full-text screening, data extraction, and writing [3]. Systematic reviews are extremely labour- and cost-intensive, taking around 67.3 weeks to complete [4], and reviewing abstracts alone takes around 40 continuous hours [4].

Because each abstract and full text is typically double-screened, the time and effort required before even writing the systematic review is extensive [5]. As research fields constantly evolve, the volume of published literature has become overwhelming to sift through. For this reason, researchers are now exploring the utility of artificial intelligence (AI) to support systematic reviews. Human error resulting from methodological choices may also add time [1]. Ensuring systematic reviews are error-free is crucial to maintaining internal validity and preventing recirculation of publication bias [1]. Using AI may reduce person-hours and help minimize bias associated with the methodological process, as it could standardize decision-making in screening [6] and data extraction [7].

Elicit.org (Elicit) is a commercially available language learning model developed by Ought, specifically marketed for systematic reviews. It uses semantic similarity to compile a list of papers in response to a research question[8]. The search database includes 115 million papers from Semantic Scholar, a separate AI-powered scientific literature search portal, as well as PDFs uploaded by the researcher [8]. The use of AI in synthesizing scientific research is controversial, with concerns about its accuracy and reliability [7], as any AI-induced biases would be detrimental to the sensitive field of healthcare [7]. Since Elicit’s formal launch in 2023, 9 studies have independently evaluated it: four [6, 9–11] evaluated search and screening, and six evaluated data extraction [11–16]. Current evaluations of Elicit conclude that while it is promising, human oversight is still necessary. Elicit’s sensitivity during the screening process was found to be much lower than that of human reviewers [6, 9–11]. In terms of data extraction, Elicit achieved 77-100% accuracy in extracting population demographics [11–16], demonstrating the tool’s utility. Elicit also showed some cases of confabulation [12, 13], raising concerns about its accuracy. To our knowledge, no peer-reviewed systematic review has explicitly stated that Elicit was used for its screening stages in its methods.

Our laboratory (Shah et al.[17]) recently completed a systematic review on the topic of whether psychosis spectrum symptoms (PSS) are a risk factor for subsequent mental health diagnoses. PSS refer to subclinical psychotic symptoms (inclusive of psychotic experiences, psychotic-like experiences, attenuated disorganization, and attenuated negative symptoms) [18–20]. The way PSS is measured and reported is heterogeneous, which increases the time and labour expenses associated with investigating PSS through a systematic review. Our review therefore sought primary papers where both PSS was measured at baseline and mental health diagnoses were assessed at least a year later. These reports may have been nested within larger longitudinal investigations that also reported other variables and subsamples outside our research question’s scope. We therefore consider this research question an ideal use case for evaluating Elicit’s ability to support the systematic review process. After completing our manual review, we attempted to use Elicit to replicate this systematic review. We independently evaluated Elicit’s performance by comparing the publication lists and extracted data tables at each step of the systematic review process.

## Methods

We independently evaluated Elicit’s performance at multiple stages of the systematic review process by testing its ability to replicate an existing systematic review and meta-analysis recently published by our research group (cite Jiya paper). We compared systematic review stages according to PRISMA reporting guidelines, including study selection, title and abstract screening, full-text screening, and data extraction. After manually completing these stages, we purchased a paid (“Pro-tier”) Elicit subscription between December 2025 and May 2026, with study co-author (IW) as the primary user. IW was instructed to use the software to replicate the manual systematic review results and met weekly with the manual systematic review authors (JS and EWD) to ensure the search terms and criteria were consistent with the manual review. Finally, we calculated accuracy, precision, and error rate by comparing the studies included in the manual and Elicit reviews and the data extraction.

### Manual systematic review

We conducted the manual systematic review following PRISMA guidelines [5] using the Covidence platform [21]. Titles and abstracts were independently screened by 4 reviewers (JS, CD, YA, AM). Full-text screens were reviewed by 3 reviewers (JS, CD, IW); all abstracts and full text required inclusion by two reviewers in order to proceed to the next stage. A third reviewer resolved any discrepancies. The manual PSS systematic review aimed to investigate whether the presence of youth PSS is a risk factor for the emergence of mental health disorders. We searched Ovid Medline, PsycINFO, Web of Science, and EMBASE from inception to March 2025. The inclusion criteria are as follows:

a. Minimum 1-year follow-up period
b. The average age of participants must be 25 or below at baseline.
c. Participants in the study must be assessed for baseline PSS. PSS could have been identified through self-report questionnaires, structured research interviews, or clinical interviews. PSS is defined as experiencing any of the following: brief psychotic experiences (delusions, hallucinations, thought disorganization), persistent psychotic experiences, psychotic like experiences (attenuated delusions or perceptual abnormalities), attenuated disorganization or attenuated negative symptoms[18–20].
d. Studies must ascertain diagnostic outcomes at follow-up using standardized criteria from any edition of the Diagnostic and Statistical Manual of Mental Disorders (DSM)[22] or the International Classification of Diseases (ICD)[23] collected via established semi-structured interviews conducted by trained assessors.
e. Participants must not meet baseline threshold psychosis disorder symptoms, nor have been diagnosed with first episode psychosis, or any psychotic disorder at baseline.
f. Must be a primary research article published before August 2025 in a peer-reviewed journal, written in English (we excluded controlled clinical trials, case series, dissertations, abstracts, grey literature, and study protocols)

### Elicit study selection

Unlike traditional search engines, Elicit’s response depends on the research question posed, rather than truncation or search syntax [24]. The input question used in Elicit’s search was: “Are psychosis spectrum symptoms predictive of emergent mental health disorders diagnosed using any version of DSM or ICD across community-based populations for youth with a mean age of 25 or less, and are they associated with a number of mental health and functional outcomes using longitudinal studies with a minimum follow-up period of 1 year?”

### Elicit screening functions

At the time of the study (April 2026), Elicit could output 1000 studies using its own search strategy based on an input question, while allowing up to 500 additional studies to be supplemented by the user. To improve comparability between Elicit screening and the manual systematic review, we supplemented the Elicit initial base list with the 233 studies that reached the full-text screening stage in the manual review. We manually removed duplicates after the study selection process. We manually uploaded missing PDFs for supplemented studies for full-text screening.

The screening criteria given to Elicit are detailed in Supplemental Table 1. When screening studies, Elicit assigns each study a score from 0 to 5 based on how well it meets the screening criteria, and the threshold score determines which studies are included or excluded. Threshold settings for the abstract and full-text screening stages were both set to 2.5 out of 5, the default. The strict criterion feature, which results in an automatic exclusion if the paper failed to meet the respective criterion regardless of its study score, was used for four criteria: 1) the diagnostic outcomes (i.e. the presence of a mental health disorder diagnosis at follow-up using any version of the DSM/ICD), 2) study design follow-up time (> 1 year), 3) presence of a systematic review or poster, and 4) the publication date (i.e. published before August 2025 - matching the manual systematic review).

### Evaluating Elicit’s data extraction

The criteria for data extraction are given in Supplemental Table 2. To evaluate Elicit’s data extraction, we compared Elicit’s results with the manual review results. Matches were coded by IW for each extracted value on a scale of 0 (completely inaccurate) to 2 (exact match), where “2 (exact match)” was assigned when Elicit found an exact match to the value attained through the manual data extraction process; “1 (inaccurate but helpful)” was given if the data collected wasn’t an exact match with the manual data extraction, but had a sentence-specific citation that was helpful (i.e. if the average age wasn’t found in but Elicit had extracted the age range); “0 (completely inaccurate)” was given if there wasn’t an exact match, or any helpful citations, or if irrelevant data was noted.

### Accuracy, precision, sensitivity, and error measurements

We assessed primary outcomes across the two screening stages of Elicit’s systematic review, using the manual review results as the standard for comparison. Primary outcomes used True Negative (TN), True Positive (TP), False Negative (FN), and False Positive (FP) values to calculate accuracy, precision, sensitivity, and error rate.

### Person-time savings

The secondary outcome assessed at each stage of the systematic review process is person-time savings, defined as the total time spent on the systematic review across different people (4).

## Results

### Study selection

See Figure 1 (workflow schematic) for the comparison of Elicit and Manual workflows. Elicit’s initial search yielded 1000 studies; of these, only 158 (15.8%) overlapped with our manual review, for a precision of 0.158 (Figure 1).

**Figure 1.**
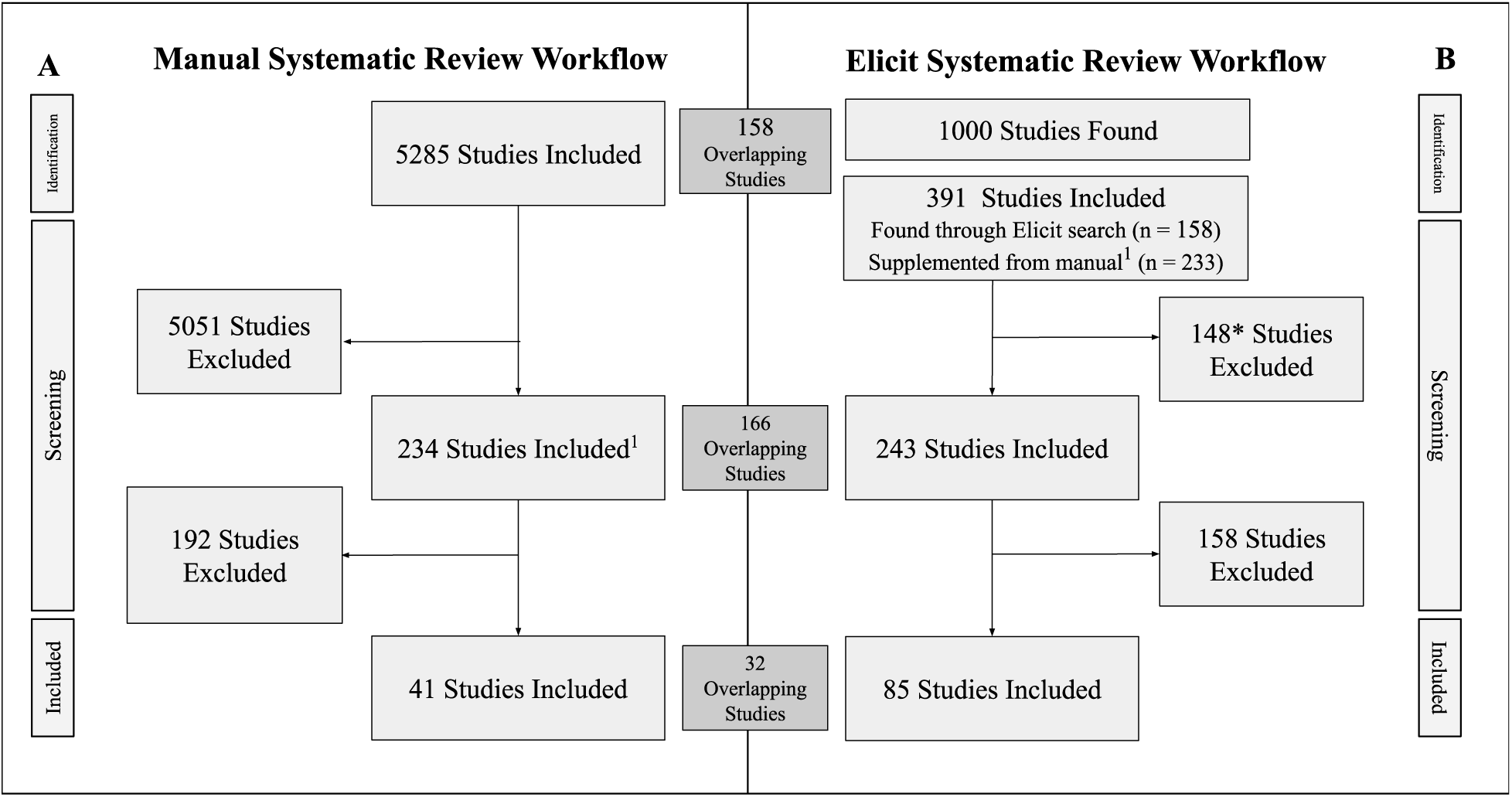
Comparison of manual and Elicit systematic review workflows: 1A depicts the manual systematic review workflow, where a team of 5 double-screened each stage. 1B depicts the Elicit systematic review workflow, where each stage was screened once by the AI tool. We evaluated abstract and full-text screening using the 391 overlapping studies found through Elicit’s search selection process and the manual search process. ^1^ = 233 studies included after abstract screening in the manual systematic review were added to Elicit’s base study list.

### Screening: Accuracy, precision, sensitivity, and error measurements

To expand the search pool for subsequent steps, we supplemented the search list with an additional 233 studies from the full-text screening list of our manual review. Of the 233 supplemented studies, 6 failed to upload, leaving 227 studies supplemented from the manual review total. To ensure that Elicit’s screening process was evaluated without the addition of studies that were not manually screened, accuracy, precision, sensitivity, and error rates were calculated with TP, TN, FP, and FN values derived from the number of studies at each stage that overlapped with the original 391 studies that overlapped with the manual review (Figure 2). For Elicit’s abstract screening stage, moderate performance was observed (accuracy 0.63, sensitivity 0.71, precision 0.68, and error rate 0.37) (Figure 2). However, 3 studies in the final manual study list were excluded at abstract screening [25–27]. For the full-text screening process, accuracy and sensitivity remained moderate (accuracy was 0.74, sensitivity was 0.78, error rate 0.26) (Figure 2), while precision was lower (0.38). With both screening steps taken together, this resulted in an overall accuracy of 0.84, with a precision of 0.38 and sensitivity of 0.78.

**Figure 2.**
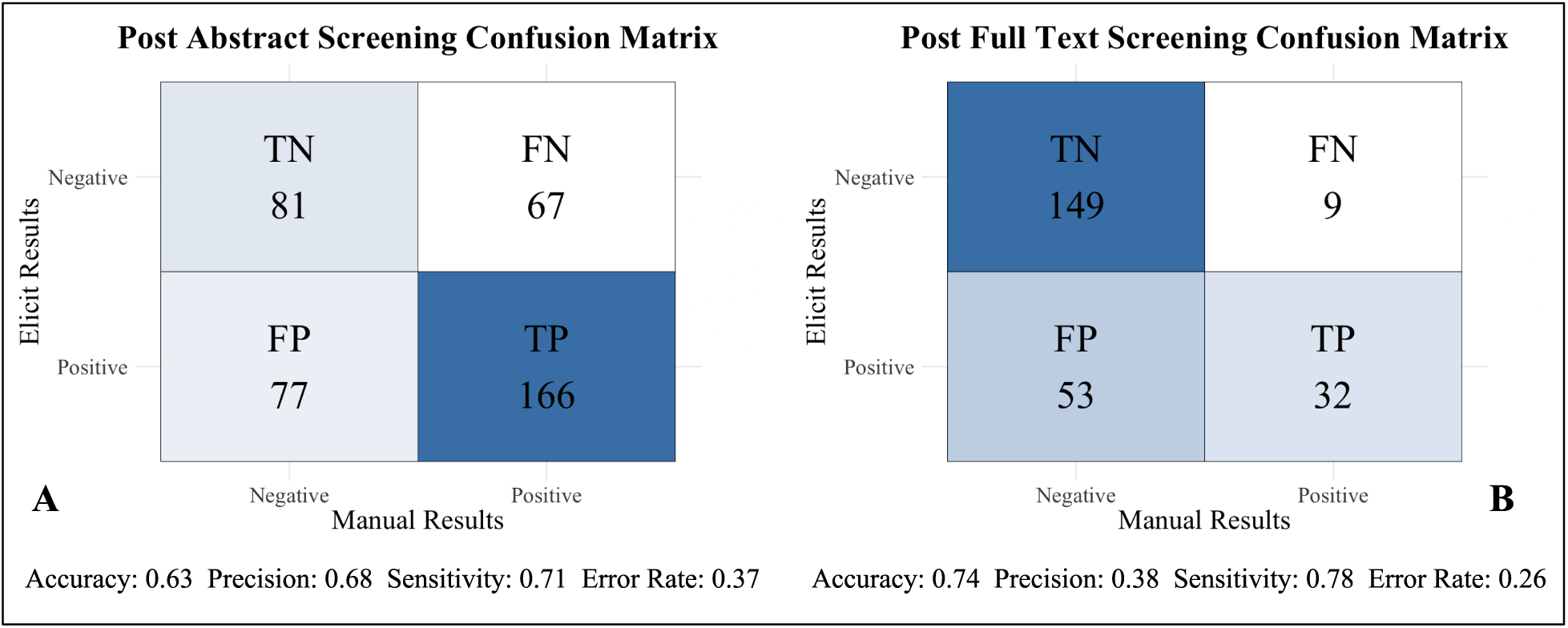
Moderate sensitivity with poor precision in Elicit’s abstract and full-text screening stages: 2A and 2 B depict confusion matrices for their respective screening stages, where TN = true negative, TP = true positive, FP = false positive, and FN = false negative. 2A depicts a confusion matrix of Elicit’s post-abstracts screening stage. 2B depicts a confusion matrix of Elicit’s post-full-text screening stage. 2AB were both coded in RStudio using v. 4.5.2.

Figure 3 summarizes screening errors at the full-text screening stage. For false negatives, Elicit falsely excluded six studies for having incorrect diagnostic outcomes (Figure 3). Diagnostic outcomes referred to the presence of a mental health disorder diagnosis at follow-up using any version of the DSM/ICD. Elicit excluded one study for lacking the full text despite having the full-text PDF manually uploaded. Elicit also falsely excluded one study because it scored below Elicit’s default inclusion threshold of 2.5.

**Figure 3.**
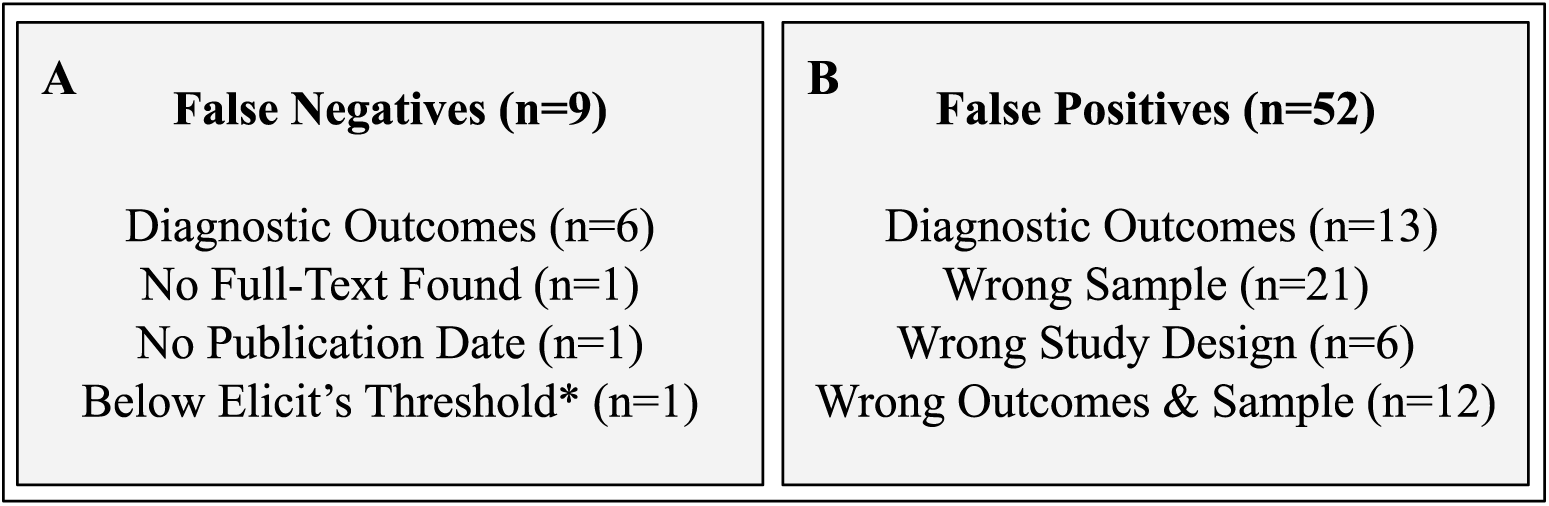
False negatives and false positives characterization at the full-text screening stage: 3A depicts Elicit’s cited reason for exclusion for each study. 3B showcases the reasons why the incorrectly included studies should have been excluded. The characterization of No Full-Text Found in 3AB refers to any scenario where the study was a poster, oral abstract, conference abstract, or where no corresponding full-text paper was found.

For false positives, many should have been excluded because they analyzed the wrong sample or had incorrect diagnostic outcomes. Regarding diagnostic outcomes, many false-positive studies used worsening symptoms as the outcome instead of a diagnosis. For studies with “Wrong Study Design,” there were three conference abstracts, two posters, and one letter to the editor included after full-text screening.

### Accuracy of Elicit’s data extraction

In total, 34 of 41 studies were evaluated for data extraction, as 2 studies failed to upload and 5 did not report enough data for extraction. For data extraction, “perfect matches” were extracted in over 85% of cases for author, year, country, PSS assessment used, and effect type (*Tab. 1*). Even when a perfect match was not extracted, useful information was extracted in >85% of cases for follow-up time (*Tab. 2*). The effect (i.e. extraction of the test statistic for meta-analysis), average age, and female/male ratio, were the criteria with the lowest “perfect matches” (*Tab. 1*). For sample age and sample female/male ratio, most errors stemmed from extracting these statistics from the incorrect subsample within a paper that reported on multiple subgroups or timepoints. In addition, Elicit typically reported only the total follow-up time in cases, or all follow-up times in cases with multiple follow-up times. Effect was the criterion with the lowest proportion of data extractions offering either a “perfect match” or useful information.

**Table 1:** Comparing Elicit data extraction tool to manual extraction (k = 34 studies) The table shows the proportion of studies (0 to 1) whose data extraction from Elicit matched the data extracted in the manual systematic review. The second column shows the proportion of studies meeting the 1-threshold (scores of 1 or above). The third column shows the proportion of studies meeting the 2-threshold, including studies scoring 2 or higher. All 41 studies included in the manual systematic review were fed through Elicit’s data extraction feature. Data from 34 studies were analyzed due to some studies in the manual review not having enough data for analysis.

| **Criterion** | **Proportion meeting exact criterion** | **Proportion offering helpful information for criterion** | **Description of Elicit's extraction errors** |
| --- | --- | --- | --- |
| **Author** | 0.97 | 0.97 | No author information was extracted[28]. |
| **Year** | 0.94 | 0.94 | Original publishing date was the year before, being the EPUB date. [29, 30] |
| **Average age** | 0.61 | 0.82 | Elicit provided an inaccurate age range [30, 31]<br>Age range given, but no average age [32–37]<br>Average age found was from a follow-up time rather than at baseline [38, 39] |
| **Follow-up time** | 0.68 | 0.91 | Follow-up time given by Elicit was incorrect [28, 40]<br>Follow-up time only included the<br>total time, and not the time from when the first PSS were assessed [30, 37, 41–43]<br>When follow-up time was variable, manual reviewers chose a different value than Elicit. [26, 44–46] |
| **Sample size** | 0.76 | 0.82 | Multiple sample sizes were used, and the sample size Elicit chose wasn't at baseline [36].<br>Sample size listed was not given in the paper [31, 47–49].<br>Elicit did not include one or more of the groups in their sample size [29, 34, 50]. |
| **Female/male ratio** | 0.62 | 0.71 | Elicit did not include the ratio despite it being listed, sometimes in a table [32, 34–36, 38, 48, 49].<br>Elicit calculated the ratio using the percentage of boys from a base sample, but the base sample used differed from the total sample size [30, 42, 51]. |
| **Country** | 0.91 | 0.91 | Elicit did not extract country information [52].<br>Elicit extracted different country information [40, 44]. |
| **PSS assessment used** | 0.88 | 0.88 | Elicit did not extract PSS assessment information [29]<br>Elicit extracted a different PSS assessment [30, 41, 53]. |
| **Effect type** | 0.88 | 0.88 | No information on defining the effect type was extracted [29, 44].<br>An effect type was identified, but it wasn't for the specific effect required [31, 52, 54, 55]. |
| **Effect** | 0.65 | 0.65 | The correct effect type is identified, but the effect itself is either incorrect or absent [28, 30, 31, 35, 41, 43, 56, 57]. |
| **<u>All criteria</u>** | **<u>0.79</u>** | **<u>0.85</u>** |  |

**Table 2:** Elicit’s screening and extraction processes take less time. Person-time spent at each stage of both reviews. The time calculated for the manual systematic review includes five people for title & abstract screening, three people for full-text screening, and one person for data extraction. Prep at each stage refers to any troubleshooting that occurred before or during each stage.

|  | **Time for each manual systematic review step (hrs)** | **Time for each Elicit systematic review step (hrs)** |
| --- | --- | --- |
| **Prep** | 2 | 0.33 |
| **Title & abstract** | 352.3 | 0.22 |
| **Full text prep** | 0 | 2 |
| **Full text** | 78 | 0.5 |
| **Data extraction** | 12.67 | 0.12 |
| **Total time** | 444.97 | 3.16 |

### Person-time savings

See Table 2 for comparison of the person-time spent at each stage of the systematic review screening and data extraction process. Elicit required much less time. The total person-time for the manual review was 453.97 hours, and the total person-time for the Elicit review was 3.16 hours.

## Discussion

This study evaluated the AI tool Elicit’s ability to conduct the screening and data extraction steps of a systematic review by comparing its results at each screening stage with the manual review. Elicit’s search was poor, with only 158 out of 1000 studies overlapping with the 5285 studies of the manual review. During both abstract and full-text screening stages, Elicit had moderate precision and high sensitivity. In comparison, 79% of all data extracted during the data extraction step matched the data extracted from the manual review, with straightforward categories like author, country, and year having many exact matches. In terms of person-time savings, Elicit is a lot less time-intensive to run, with its entire systematic review process taking 3.16 hours. Though Elicit shows promise as an adjunct screening and data extraction tool, it is not ready in its current form to act as a standalone tool.

### Elicit’s study selection

Using semantic search, Elicit found very few overlapping studies with the ones found through traditional search methods. At the time of this writing, Elicit limits search results to 1000 studies using its semantic search at the “pro user” function. This meant it would never fully replicate the 5285-study list generated for our original manual review. Because many systematic reviews in our field start with a study list well over 1000, this is a serious challenge to the platform’s utility. Of the 1000 studies Elicit found, 158 overlapped with the 5285 initially found using the manual review’s search strategy. The low number of overlapping studies is concerning. This finding replicates previous reports, where the number of overlapping studies to published reviews ranges from 8% [10] to 50% [11]. This may be because fewer studies that might have been included could not be imported. One possible explanation for this discrepancy is the difference in source databases. However, we confirmed that all studies included in the final manual review were available in Semantic Scholar. Still, 27/41 (66%) of the studies included in our final meta-analysis were not returned through Elicit’s search function. This leaves a strong divergence between semantic search strategy results and those from a more traditional search strategy.

### Elicit’s abstract and full-text screening is modest, but biased

In both the Abstract and Full-Text Screening Stages, Elicit showed moderate accuracy (74% after full text) with poor precision (37%) and moderate sensitivity (78%), with a skew toward false positives. These values align with previous studies [6, 9, 10], which concluded that Elicit is not ready for broader use. Though we gave Elicit a difficult use case, major false-positive errors such as including posters, abstracts, and non-peer-reviewed preprints still should not have occurred. A skew toward false positives may be preferred for a “human-teamed” screening system, where more human time dealing with false positives may be preferred to excluding relevant studies. Therefore, the less-than-perfect sensitivity, or the presence of false negatives, is the most concerning aspect of Elicit, as should Elicit been used solely to screen papers for the manual review, 21% of the papers that made it into manual review would have been excluded. In our case, the majority of falsely excluded studies were excluded for lacking the correct diagnostic outcomes, meaning that Elicit failed to find evidence that the paper reported a diagnostic outcome using any version of the DSM/ICD, when manual checks confirmed that the studies had.

In general, we observed a skew toward false positives. Note that a skew toward false positives in difficult cases may have been expected, as we used a rather liberal threshold setting of 2.5 (the Elicit default). We also found that Elicit tends to err on the side of caution when excluding studies, especially when unclear whether a study fits the evaluation criteria perfectly. These false positives have been previously reported. For example, Lau and Golder [9] also noted that Elicit would include papers published outside their specified date range. However, Lau and Golder’s (2025) study was conducted before the release of the strict criterion feature. We observed that the use of strict criterion does not remove all false positives at the full-text stage. Note that even with this liberal threshold, we did not achieve perfect sensitivity; we also observed false negatives.

### Data extraction

Although the Elicit data extraction tool’s results were imperfect, the authors agreed that the interface was very helpful to use. For the overall criteria match in data extraction, 85% of the data that Elicit found was considered helpful to the human user (“helpful”), and 79% of the data found was fully accurate (“accurate”). These accuracies are in agreement with previous studies [11–16], where Elicit has outperformed other AI tools [11, 13, 15, 16], but failed to equal manual human extractors [16]. The criteria with the greatest proportion of accurate and helpful matches are author, year, and country; these are also straightforward categories that typically can be extracted from file metadata and don’t require a deep search. Many other data extraction tools on the market may also extract data in these straightforward categories, such as Covidence (2026). Elicit was also mostly accurate in identifying the PSS assessment used, which was usually embedded in each paper. Elicit’s greatest asset in data extraction is its sentence-specific citations, which allow Elicit to cite the sentence from the original paper from which they pulled the data. However, human oversight is still needed for data extraction. Some parts of the data extraction that were difficult for manual reviewers to extract were also difficult for Elicit to extract. The criterion “effect,” which refers to the category of hazard ratios (HR), odds ratios (OR), or risk ratios (RR) reported and the corresponding ratio, had the lowest proportion of correct matches to the manual review. Upon examination, this may be because some studies report multiple HRs, ORs, and RRs, and Elicit’s data extraction may not extract the effect or may extract the wrong effect. Importantly, “effect” statistics are critical for meta-analytic statistical calculations; therefore, failing to extract these values would prevent us from reproducing our meta-analytic results. In our use case, extraction errors were largely due to picking the “wrong” statistic within a paper where multiple sub-analyses (or sub-samples) were reported on. This difficult case is common for research questions such as ours, where we aimed to synthesize a result that may have been a secondary aim in multiple primary papers.

### Elicit is less time- and cost-intensive to run

In terms of person-time, Elicit is less time- and cost-intensive to run, taking 3.16 hours to complete most of the systematic review process compared to the 444.97 total hours required for the manual systematic review. Additionally, the monthly subscription to Elicit was $42 USD, which is less costly than the labour costs of five reviewers for the manual systematic review. As a result, Elicit could reduce the time needed for systematic reviews, especially for data extraction, while remaining cost-effective. Even when Elicit wasn’t fully accurate, its sentence-specific citations could increase reviewer efficiency and potentially change how double screening is understood for data extraction. Instead of having two human reviewers, there could be one reviewer who oversees Elicit’s data extraction to ensure that the data is correctly extracted correctly.

There is a lot of troubleshooting involved in tailoring Elicit’s responses. Despite Elicit’s access to over 250 million papers through databases including PubMed, Semantic Scholar, and OpenAlex (2025), most papers pulled up through Elicit’s search process did not automatically upload the full-text PDF. Some papers lacked a PDF upload because they were conference abstracts or e-posters without an associated full text, while others simply lacked the full text. As a result, users still needed to manually upload the missing PDFs, which accounted for most of the person-time spent on the Elicit systematic review process. Despite this, the time needed to troubleshoot Elicit was still significantly shorter than the time taken for any step of the manual systematic review, which still makes Elicit the more time-efficient option.

### Strengths and limitations

As this project was completed between December 2025 and April 2026, we acknowledge that there have been updates made to Elicit, and that some of the errors and issues we encountered during troubleshooting may be resolved. This paper treats manual review as the standard for comparison and does not account for inter-human error. For a thorough comparison of Elicit performance, we could have evaluated it against the performance of an independent human team. Although two human reviewers independently reviewed each study, with a third reviewer in some cases, inter-human error may still have occurred. Only one user used Elicit so that an error may have occurred unintentionally. Future studies could evaluate Elicit with multiple users and multiple runs.

### Conclusions

Overall, Elicit is promising as an adjunct screening and data extraction tool. However, it is not yet ready for use as a standalone tool. Elicit’s search function was poor, with only 16% of studies overlapping with the studies found using the manual search strategy. Elicit’s screening stages were modest, with both stages being skewed towards false positives. Though we acknowledge that we gave Elicit a difficult systematic review question, the appearance of major errors, like including abstracts and posters after full-text screening, is not understandable. Elicit’s data extraction interface was also user-friendly, and its sentence-specific citations could help increase person-time savings. However, Elicit’s data extraction is less promising for more nuanced criteria, such as follow-up time in cases with multiple follow-ups. Rather than using Elicit as the sole screening or data extraction tool, we recommend treating it as a second or third reviewer, with conflicts between Elicit and a manual reviewer resolved by a third human reviewer. Future evaluations could also examine person-time savings and accuracy when using Elicit in this capacity.

## Data Availability

All data produced in the present study are available upon reasonable request to the authors

## Acknowledgements

We thank the four reviewers who helped screen titles and abstracts, as well as full texts, for the manual systematic review: Jiya Shah and Clarize Donato. Anisha Miah and Yar Al Dabagh. This work was supported by the Canadian Institutes of Health Research - Institute for Human Development, Child and Youth Health and the University of Toronto Department of Psychiatry Academic Scholar’s Award.

## References

1. Smith KA, Cipriani A, Geddes JR. The usefulness and interpretation of systematic reviews. BJPsych Adv. 2016 Mar;22(2):132–41. doi:10.1192/apt.bp.114.013128

2. Grant MJ, Booth A. A typology of reviews: an analysis of 14 review types and associated methodologies. Health Inf Libr J. 2009;26(2):91–108. doi:10.1111/j.1471-1842.2009.00848.x

3. Motevalli M. Comparative Analysis of Systematic, Scoping, Umbrella, and Narrative Reviews in Clinical Research: Critical Considerations and Future Directions. Int J Clin Pract. 2025;2025(1):9929300. doi:10.1155/ijcp/9929300

4. Pang X, Saif-Ur-Rahman K, Berhane S, Yao X, Kothari K, Taneri PE, et al. Comparing Artificial Intelligence and manual methods in systematic review processes: protocol for a systematic review. J Clin Epidemiol. 2025 May 1;181:111738. doi:10.1016/j.jclinepi.2025.111738

5. Randles R, Finnegan A. Guidelines for writing a systematic review. Nurse Educ Today. 2023 Jun 1;125:105803. doi:10.1016/j.nedt.2023.105803

6. Brugha T.S., Morgan Z., Bebbington P., Jenkins R., Lewis G., Farrell M., et al. Social support networks and type of neurotic symptom among adults in British households. Psychol Med. 2003;33(2):307EP – 318. Located at: 36221462. doi:10.1017/S0033291702006505

7. Page MJ, Moher D, Bossuyt PM, Boutron I, Hoffmann TC, Mulrow CD, et al. PRISMA 2020 explanation and elaboration: updated guidance and exemplars for reporting systematic reviews. BMJ. 2021 Mar 29;372:n160. doi:10.1136/bmj.n160 PubMed PMID: 33781993.

8. Bernard N, Sagawa Jr Y, Bier N, Lihoreau T, Pazart L, Tannou T. Using artificial intelligence for systematic review: the example of elicit. BMC Med Res Methodol. 2025 Mar 18;25(1):75. doi:10.1186/s12874-025-02528-y

9. Ge L, Agrawal R, Singer M, Kannapiran P, De Castro Molina JA, Teow KL, et al. Leveraging artificial intelligence to enhance systematic reviews in health research: advanced tools and challenges. Syst Rev. 2024 Oct 25;13(1):269. doi:10.1186/s13643-024-02682-2

10. Elicit: AI literature review research assistant. Public Serv Q. 2023;19(3):201–7. doi:10.1080/15228959.2023.2224125

11. Kung J. Elicit (product review). J Can Health Libr Assoc. 2023 Apr 1;44(1). doi:10.29173/jchla29657

12. Helms Andersen T, Marcussen TM, Termannsen AD, Lawaetz TWH, Nørgaard O. Using Artificial Intelligence Tools as Second Reviewers for Data Extraction in Systematic Reviews: A Performance Comparison of Two AI Tools Against Human Reviewers. Cochrane Evid Synth Methods. 2025;3(4):e70036. doi:10.1002/cesm.70036

13. Hupe M. Covidence. J Electron Resour Med Libr. 2025 Jul 3;22(3):139–47. doi:10.1080/15424065.2025.2547027

14. Welham J, Scott J, Williams G, Najman J, Bor W, O’Callaghan M, et al. Emotional and behavioural antecedents of young adults who screen positive for non-affective psychosis: a 21-year birth cohort study. Psychol Med. 2009;39(4):625–34. Located at: WOS:000264691000009. doi:10.1017/S0033291708003760

15. Coughlan H, Healy C, Ni Sheaghdha A, Murray G, Humphries N, Clarke M, et al. Early risk and protective factors and young adult outcomes in a longitudinal sample of young people with a history of psychotic-like experiences. Early Interv Psychiatry. 2020;14(3):307–20. doi:10.1111/eip.12855

16. Stanfield AC, McKechanie AG, Lawrie SM, Johnstone EC, Owens DGC. Predictors of psychotic symptoms among young people with special educational needs. Br J Psychiatry. 2019;215(1):422–7. doi:10.1192/bjp.2018.296

17. Lindgren M., Manninen M., Kalska H., Mustonen U., Laajasalo T., Moilanen K., et al. Predicting psychosis in a general adolescent psychiatric sample. Schizophr Res. 2014;153(SUPPL. 1):S171. Located at: 71728785

18. Addington J, Liu L, Farris M, Goldstein B, Wang J, Kennedy S, et al. Clinical staging for youth at-risk for serious mental illness: A longitudinal perspective. EARLY Interv PSYCHIATRY. 2021;15(5):1188–96. Located at: WOS:000577939100001. doi:10.1111/eip.13062

19. Ärmänen A, Lahti M, Therman S, Suvisaari J, Lindgren M. Psychological, social and role functioning as predictors of psychosis in an adolescent psychiatric sample. EARLY Interv PSYCHIATRY. 2018;12(6):1064–71. Located at: WOS:000450650300006. doi:10.1111/eip.12414

20. Palomaki J, Therman S, Kerkela M, Jarvelin MR, Jones P, Murray GK, et al. Specific adolescent prodromal symptoms associated with onset of psychosis in the northern finland birth cohort 1986. Early Interv Psychiatry. 2022;(Anthony, J. C., Folstein, M., Romanoski, A. J., Von Korff, M. R., Nestadt, G. R., Chahal, R., Merchant, A., Brown, C. H., Shapiro, S., Kramer, M., Gruenberg, E. M. (1985). Comparison of the lay diagnostic interview schedule and a standardized psychiatri):No-Specified. doi:10.1111/eip.13363

21. Coughlan H, Walton-Ball E, Carey E, Healy C, O’Regan-Murphy G, Uidhir A, et al. Self-reported interpersonal and educational/vocational difficulties in young adults with a history of transient psychotic experiences: findings from a population-based study. BMC PSYCHIATRY. 2021;21(1). Located at: WOS:000609423500006. doi:10.1186/s12888-020-03022-z

22. Calkins ME, Moore TM, Satterthwaite TD, Wolf DH, Turetsky BI, Roalf DR, et al. Persistence of psychosis spectrum symptoms in the Philadelphia Neurodevelopmental Cohort: A prospective two-year follow-up. World Psychiatry. 2017;16(1):62–76. doi:10.1002/wps.20386

23. Calkins M.E., Butler E.R., Moore T.M., Ered A., Taylor J.H., White L.K., et al. Longitudinal Trajectories of Clinical Features in Community Youth with Recurrent Psychosis Spectrum Symptoms: Findings from the Philadelphia Neurodevelopmental Cohort. Schizophr Bull. 2025;51(4):858EP – 870. Located at: 2039562610. doi:10.1093/schbul/sbaf075

24. Mamah D, Musau A, Mutiso V, Owoso A, Ben Abdallah A, Cottler L, et al. Characterizing psychosis risk traits in Africa: A longitudinal study of Kenyan adolescents. Schizophr Res. 2016;176(2–3):340–8. Located at: WOS:000384130200041. doi:10.1016/j.schres.2016.08.004

25. Dhossche D, Ferdinand R, Van der Ende J, Hofstra M, Verhulst F. Diagnostic outcome of self-reported hallucinations in a community sample of adolescents. Psychol Med. 2002;32(4):619–27. Located at: WOS:000175640000006. doi:10.1017/S003329170200555X

26. Giocondo J, Salum G, Gadelha A, Argolo F, Simioni A, Mari J, et al. Psychotic-like Experiences and Common Mental Disorders in Childhood and Adolescence: Bidirectional and Transdiagnostic Associations in a Longitudinal Community-based Study. Schizophr Bull OPEN. 2021;2(1). Located at: WOS:001399411100001. doi:10.1093/schizbullopen/sgab028

27. Livny A., Reichenberg A., Fruchter E., Yoffe R., Goldberg S., Fenchel D., et al. A population-based longitudinal study of symptoms and signs before the onset of psychosis. Am J Psychiatry. 2018;175(4):351EP – 358. Located at: 621542310. doi:10.1176/appi.ajp.2017.16121384

28. Gregersen M, Jepsen J, Rohd S, Sondergaard A, Brandt J, Ellersgaard D, et al. Developmental Pathways and Clinical Outcomes of Early Childhood Psychotic Experiences in Preadolescent Children at Familial High Risk of Schizophrenia or Bipolar Disorder: A Prospective, Longitudinal Cohort Study- The Danish High Risk and Resilience Study. Am J PSYCHIATRY. 2022;179(9):628–39. Located at: WOS:000878127800011. doi:10.1176/appi.ajp.21101076

29. Carey E, Gillan D, Healy C, Dooley N, Campbell D, McGrane J, et al. Early adult mental health, functional and neuropsychological outcomes of young people who have reported psychotic experiences: a 10-year longitudinal study. Psychol Med. 2021;51(11):1861–9. Located at: WOS:000686603400013. doi:10.1017/S0033291720000616

30. Haining K., Gajwani R., Gross J., Gumley A.I., Lawrie S.M., Schultze-Lutter F., et al. Clinical and Functional Outcomes of Community-Recruited Individuals at Clinical High-Risk for Psychosis: Results from the Youth Mental Health Risk and Resilience Study (YouR-Study). Schizophr Bull Open. 2024;5(1):sgae029. Located at: 2035991018. doi:10.1093/schizbullopen/sgae029

31. Rössler W, Hengartner M, Ajdacic-Gross V, Haker H, Gamma A, Angst J. Sub-clinical psychosis symptoms in young adults are risk factors for subsequent common mental disorders. Schizophr Res. 2011;131(1–3):18–23. Located at: WOS:000295111400004. doi:10.1016/j.schres.2011.06.019

32. Hengartner M, Heekeren K, Dvorsky D, Walitza S, Rössler W, Theodoridou A. Course of psychotic symptoms, depression and global functioning in persons at clinical high risk of psychosis: Results of a longitudinal observation study over three years focusing on both converters and non-converters. Schizophr Res. 2017;189:19–26. Located at: WOS:000416392400005. doi:10.1016/j.schres.2017.01.040

33. Kieseppä V, Lang U, Healy C, O’Hare K, Díaz-Caneja C, Gülöksüz S, et al. Do psychotic symptoms predict future psychotic disorders in adolescent psychiatry inpatients? A 17-year cohort study. Psychol Med. 2025;55. Located at: WOS:001457743200001. doi:10.1017/S003329172500073X

34. Gooding DC, Tallent KA, Matts CW. Clinical Status of At-Risk Individuals 5 Years Later: Further Validation of the Psychometric High-Risk Strategy. J Abnorm Psychol. 2005;114(1):170–5. doi:10.1037/0021-843X.114.1.170

35. Dominguez M, Saka M, Lieb R, Wittchen H, van Os J. Early Expression of Negative/Disorganized Symptoms Predicting Psychotic Experiences and Subsequent Clinical Psychosis: A 10-Year Study. Am J PSYCHIATRY. 2010;167(9):1075–82. Located at: WOS:000281794200014. doi:10.1176/appi.ajp.2010.09060883

36. O’Hare K.J.M., Poulton R., Linscott R.J. Parent-Rated Schizotypy and Clinician-Rated Psychotic Experiences in Early Adolescence as Predictors of Schizophrenia Diagnosis by Middle Adulthood. Schizophr Bull. 2025;51((O’Hare, Linscott) Department of Psychology, University of Otago, Dunedin, New Zealand(Poulton) Dunedin Multidisciplinary Health and Development Research Unit, Department of Psychology, University of Otago, Dunedin, New Zealand):S107EP – S114. Located at: 2037920697. doi:10.1093/schbul/sbad158

37. Cannon M, Caspi A, Moffitt TE, Harrington H, Taylor A, Murray RM, et al. Evidence for early-childhood pan-developmental impairment specific to schizophreniform disorder: Results from a longitudinal birth cohort. Arch Gen Psychiatry. 2002;59(5):449–57. doi:10.1001/archpsyc.59.5.449

38. Zammit S, Kounali D, Cannon M, David A, Gunnell D, Heron J, et al. Psychotic Experiences and Psychotic Disorders at Age 18 in Relation to Psychotic Experiences at Age 12 in a Longitudinal Population-Based Cohort Study. Am J PSYCHIATRY. 2013;170(7):742–50. Located at: WOS:000321023700010. doi:10.1176/appi.ajp.2013.12060768

39. Fisher H, Caspi A, Poulton R, Meier M, Houts R, Harrington H, et al. Specificity of childhood psychotic symptoms for predicting schizophrenia by 38 years of age: a birth cohort study. Psychol Med. 2013;43(10):2077–86. Located at: WOS:000324024100007. doi:10.1017/S0033291712003091

40. Zhang W, Zhu Y, Sun M, Guo R, Wu G, Wang Z, et al. Longitudinal Trajectories of Psychotic-Like Experiences and Their Relationship to Emergent Mental Disorders Among Adolescents: A 3-Year Cohort Study. J Clin PSYCHIATRY. 2019;80(4). Located at: WOS:000489554400004. doi:10.4088/JCP.18m12437

41. Poulton R, Caspi A, Moffitt T, Cannon M, Murray R, Harrington H. Children’s self-reported psychotic symptoms and adult schizophreniform disorder -: A 15-year longitudinal study. Arch Gen PSYCHIATRY. 2000;57(11):1053–8. Located at: WOS:000165055700005. doi:10.1001/archpsyc.57.11.1053

42. IMAGEN Consortium, Maitra R, Horne C, O’Daly O, Papanastasiou E, Gaser C, et al. Psychotic Like Experiences in Healthy Adolescents are Underpinned by Lower Fronto-Temporal Cortical Gyrification: a Study from the IMAGEN Consortium. Schizophr Bull. 2023;49(2):309–18. Located at: WOS:000866648900001. doi:10.1093/schbul/sbac132

43. Eack SM, Prasad KMR, Montrose DM, Goradia DD, Dworakowski D, Miewald J, et al. An integrated psychobiological predictive model of emergent psychopathology among young relatives at risk for schizophrenia. Prog Neuropsychopharmacol Biol Psychiatry. 2008;32(8):1873–8. doi:10.1016/j.pnpbp.2008.08.024

44. Mustonen A, Niemela S, Nordstrom T, Murray GK, Maki P, Jaaskelainen E, et al. Adolescent cannabis use, baseline prodromal symptoms and the risk of psychosis. Br J Psychiatry. 2018;212(4):227–33. doi:10.1192/bjp.2017.52

45. Isaksson J, Vadlin S, Olofsdotter S, Aslund C, Nilsson KW. Psychotic-like experiences during early adolescence predict symptoms of depression, anxiety, and conduct problems three years later: A community-based study. Schizophr Res. 2020;215(Aleman, A., Kahn, R.S., Selten, J.P. (2003). Sex differences in the risk of schizophrenia: evidence from meta-analysis. Arch. Gen. Psychiatr. 60(6), 565-571. https://pubmed.ncbi.nlm.nih.gov/12796219 https://dx.doi.org/10.1001/archpsyc.60.6.5652003-0607):190–6. doi:10.1016/j.schres.2019.10.033

46. Denissoff A, Mustonen A, Miettunen J, Alakokkare A, Veijola J, Scott J, et al. Trajectories of adolescent psychotic-like experiences and early cannabis exposure: Results from a Finnish Birth Cohort Study. Schizophr Res. 2022;246:95–102. Located at: WOS:000822726100001. doi:10.1016/j.schres.2022.06.014

47. Rimvall M, van Os J, Verhulst F, Wolf R, Larsen J, Clemmensen L, et al. Mental Health Service Use and Psychopharmacological Treatment Following Psychotic Experiences in Preadolescence. Am J PSYCHIATRY. 2020;177(4):318–26. Located at: WOS:000537831300008. doi:10.1176/appi.ajp.2019.19070724

## References

1. Smith KA, Cipriani A, Geddes JR. The usefulness and interpretation of systematic reviews. BJPsych Adv. 2016;22:132–41.

2. Grant MJ, Booth A. A typology of reviews: an analysis of 14 review types and associated methodologies. Health Info Libr J. 2009;26:91–108.

3. Randles R, Finnegan A. Guidelines for writing a systematic review. Nurse Educ Today. 2023;125:105803.

4. Pang X, Saif-Ur-Rahman KM, Berhane S, Yao X, Kothari K, Taneri PE, et al. Comparing Artificial Intelligence and manual methods in systematic review processes: protocol for a systematic review. J Clin Epidemiol. 2025;181:111738.

5. Page MJ, Moher D, Bossuyt PM, Boutron I, Hoffmann TC, Mulrow CD, et al. PRISMA 2020 explanation and elaboration: updated guidance and exemplars for reporting systematic reviews. BMJ. 2021;372:n160.

6. Bernard N, Sagawa Y Jr, Bier N, Lihoreau T, Pazart L, Tannou T. Using artificial intelligence for systematic review: the example of elicit. BMC Med Res Methodol. 2025;25:75.

7. Ge L, Agrawal R, Singer M, Kannapiran P, Castro Molina D, Teow JA. Leveraging artificial intelligence to enhance systematic reviews in health research: advanced tools and challenges. Syst Rev. 2024;13. 10.1186/s13643-024-02682-2.

8. AI literature review research assistant. 2023;19:201–7.

9. Lau O, Golder S. Comparison of Elicit AI and traditional literature searching in evidence syntheses using four case studies. Cochrane Evid Synth Methods. 2025;3:e70050.

10. Mazzali C, Pinciroli T, Valetto MR, Dri P, Russo AG, Air and Health Atlas Study Group. Evaluating Elicit’s systematic reviews workflow in an umbrella review on air pollution and acute lower respiratory infections: a methodological study for quality appraisal. BMC Med Res Methodol. 2026;26:167.

11. Meliante LA, Coco G, Rabiolo A, De Cillà S, Manni G. Evaluation of AI tools versus the PRISMA method for literature search, data extraction, and study composition in glaucoma systematic reviews: Content analysis. JMIR AI. 2025;4:e68592.

12. Bianchi J, Hirt J, Vogt M, Vetsch J. Data extractions using a large language model (Elicit) and human reviewers in randomized controlled trials: A systematic comparison. Cochrane Evid Synth Methods. 2025;3:e70033.

13. Helms Andersen T, Marcussen TM, Termannsen AD, Lawaetz TWH, Nørgaard O. Using artificial intelligence tools as second reviewers for data extraction in systematic reviews: A performance comparison of two AI tools against human reviewers. Cochrane Evid Synth Methods. 2025;3:e70036.

14. Lagisz M, Mizuno A, Morrison K, Pollo P, Ricolfi L, Yang Y, et al. Using Elicit AI research assistant for data extraction in systematic reviews: A feasibility study across environmental and life sciences. Res Synth Methods. 2026;:1–18.

15. Spillias S, Ollerhead KM, Andreotta M, Annand-Jones R, Boschetti F, Duggan J, et al. Evaluating generative AI for qualitative data extraction in community-based fisheries management literature. Environ Evid. 2025;14:9.

16. Hilkenmeier F, Pelzer M, Stierle C, Fink-Lamotte J. Evaluating the AI tool “Elicit” as a semi-automated second reviewer for data extraction in systematic reviews: A proof-of-concept. Soc Sci Comput Rev. 2025. 10.1177/08944393251404052.

17. Shah JN, Ameis SH, Donato CA, Wei I, Miah A, Al Dabagh Y, et al. Systematic review and meta-analysis: Do youth-reported psychosis symptoms predict later mental health diagnosis? medRxiv. 2026;:2026.07. 13.26357957. 10.64898/2026.07.13.26357957.

18. Calkins ME, Moore TM, Merikangas KR, Burstein M, Satterthwaite TD, Bilker WB, et al. The psychosis spectrum in a young U.S. community sample: findings from the Philadelphia Neurodevelopmental Cohort. World Psychiatry. 2014;13:296–305.

19. Binbay T, Drukker M, Elbi H, Tanık FA, Özkınay F, Onay H, et al. Testing the psychosis continuum: differential impact of genetic and nongenetic risk factors and comorbid psychopathology across the entire spectrum of psychosis. Schizophr Bull. 2012;38:992–1002.

20. Karcher NR, Barch DM, Avenevoli S, Savill M, Huber RS, Simon TJ, et al. Assessment of the Prodromal Questionnaire-Brief Child Version for measurement of self-reported psychoticlike experiences in childhood. JAMA Psychiatry. 2018;75:853–61.

21. Covidence systematic review software, Veritas Health Innovation. Melbourne, Australia.

22. American Psychiatric Association. Diagnostic and Statistical Manual of Mental Disorders (DSM-5 (R)). American Psychiatric Association Publishing; 2013.

23. ICD-11 for Mortality and Morbidity Statistics. https://icd.who.int/browse/2026-01/mms/en. Accessed 29 Sept 2026.

24. Kung J. Elicit (product review). J Can Health Libr Assoc. 2023;44. 10.29173/jchla29657.

25. Welham J, Scott J, Williams G, Najman J, Bor W, O’Callaghan M, et al. Emotional and behavioural antecedents of young adults who screen positive for non-affective psychosis: a 21-year birth cohort study. Psychol Med. 2009;39:625–34.

26. Coughlan H, Healy C, Ní Sheaghdha Á, Murray G, Humphries N, Clarke M, et al. Early risk and protective factors and young adult outcomes in a longitudinal sample of young people with a history of psychotic-like experiences. Early Interv Psychiatry. 2020;14:307–20.

27. Stanfield AC, McKechanie AG, Lawrie SM, Johnstone EC, Owens DGC. Predictors of psychotic symptoms among young people with special educational needs. Br J Psychiatry. 2019;215:422–7.

28. Lindgren M, Manninen M, Kalska H, Mustonen U, Laajasalo T, Moilanen K, et al. Predicting psychosis in a general adolescent psychiatric sample. Schizophr Res. 2014;158:1–6.

29. Addington J, Liu L, Farris MS, Goldstein BI, Wang JL, Kennedy SH, et al. Clinical staging for youth at-risk for serious mental illness: A longitudinal perspective. Early Interv Psychiatry. 2021;15:1188–96.

30. Ärmänen A, Lahti M, Therman S, Suvisaari J, Lindgren M. Psychological, social and role functioning as predictors of psychosis in an adolescent psychiatric sample. Early Interv Psychiatry. 2018;12:1064–71.

31. Palomäki J, Therman S, Kerkelä M, Järvelin M-R, Jones P, Murray GK, et al. Specific adolescent prodromal symptoms associated with onset of psychosis in the Northern Finland Birth Cohort 1986. Early Interv Psychiatry. 2023;17:692–701.

32. Calkins ME, Moore TM, Satterthwaite TD, Wolf DH, Turetsky BI, Roalf DR, et al. Persistence of psychosis spectrum symptoms in the Philadelphia Neurodevelopmental Cohort: a prospective two-year follow-up. World Psychiatry. 2017;16:62–76.

33. Calkins ME, Butler ER, Moore TM, Ered A, Taylor JH, White LK, et al. Longitudinal trajectories of clinical features in community youth with recurrent psychosis spectrum symptoms: Findings from the Philadelphia Neurodevelopmental Cohort. Schizophr Bull. 2025;51:858–70.

34. Mamah D, Musau A, Mutiso VN, Owoso A, Abdallah AB, Cottler LB, et al. Characterizing psychosis risk traits in Africa: A longitudinal study of Kenyan adolescents. Schizophr Res. 2016;176:340–8.

35. Dhossche D, Ferdinand R, Van der Ende J, Hofstra MB, Verhulst F. Diagnostic outcome of self-reported hallucinations in a community sample of adolescents. Psychol Med. 2002;32:619–27.

36. Giocondo JG, Salum GA, Gadelha A, Argolo FC, Simioni AR, Mari JJ, et al. Psychotic-like Experiences and Common Mental Disorders in childhood and adolescence: Bidirectional and transdiagnostic associations in a longitudinal community-based study. Schizophr Bull Open. 2021;2. 10.1093/schizbullopen/sgab028.

37. Livny A, Reichenberg A, Fruchter E, Yoffe R, Goldberg S, Fenchel D, et al. A population-based longitudinal study of symptoms and signs before the onset of psychosis. Am J Psychiatry. 2018;175:351–8.

38. Gregersen M, Jepsen J, Rohd S, Sondergaard A, Brandt J, Ellersgaard D. Developmental Pathways and Clinical Outcomes of Early Childhood Psychotic Experiences in Preadolescent Children at Familial High Risk of Schizophrenia or Bipolar Disorder: A Prospective, Longitudinal Cohort Study- The Danish High Risk and Resilience Study. Am J PSYCHIATRY. 2022;179:628–39.

39. Carey E, Gillan D, Healy C, Dooley N, Campbell D, McGrane J, et al. Early adult mental health, functional and neuropsychological outcomes of young people who have reported psychotic experiences: a 10-year longitudinal study. Psychol Med. 2021;51:1861–9.

40. Haining K, Gajwani R, Gross J, Gumley AI, Lawrie SM, Schultze-Lutter F, et al. Clinical and functional outcomes of community-recruited individuals at clinical high-risk for psychosis: Results from the Youth Mental Health Risk and Resilience Study (YouR-Study). Schizophr Bull Open. 2024;5:sgae029.

41. Rössler W, Hengartner MP, Ajdacic-Gross V, Haker H, Gamma A, Angst J. Sub-clinical psychosis symptoms in young adults are risk factors for subsequent common mental disorders. Schizophr Res. 2011;131:18–23.

42. Hengartner MP, Heekeren K, Dvorsky D, Walitza S, Rössler W, Theodoridou A. Course of psychotic symptoms, depression and global functioning in persons at clinical high risk of psychosis: Results of a longitudinal observation study over three years focusing on both converters and non-converters. Schizophr Res. 2017;189:19–26.

43. Kieseppä V, Lång U, Healy C, O’Hare K, Díaz-Caneja CM, Gülöksüz S, et al. Do psychotic symptoms predict future psychotic disorders in adolescent psychiatry inpatients? A 17-year cohort study. Psychol Med. 2025;55:e108.

44. Gooding DC, Tallent KA, Matts CW. Clinical status of at-risk individuals 5 years later: further validation of the psychometric high-risk strategy. J Abnorm Psychol. 2005;114:170–5.

45. Dominguez M-G, Saka MC, Lieb R, Wittchen H-U, van Os J. Early expression of negative/disorganized symptoms predicting psychotic experiences and subsequent clinical psychosis: a 10-year study. Am J Psychiatry. 2010;167:1075–82.

46. O’Hare KJM, Poulton R, Linscott RJ. Parent-rated schizotypy and clinician-rated psychotic experiences in early adolescence as predictors of schizophrenia diagnosis by middle adulthood. Schizophr Bull. 2025;51 Suppl 2:S107–14.

47. Cannon M, Caspi A, Moffitt TE, Harrington H, Taylor A, Murray RM, et al. Evidence for early-childhood, pan-developmental impairment specific to schizophreniform disorder: results from a longitudinal birth cohort. Arch Gen Psychiatry. 2002;59:449–56.

48. Zammit S, Kounali D, Cannon M, David AS, Gunnell D, Heron J, et al. Psychotic experiences and psychotic disorders at age 18 in relation to psychotic experiences at age 12 in a longitudinal population-based cohort study. Am J Psychiatry. 2013;170:742–50.

49. Fisher H, Caspi A, Poulton R, Meier M, Houts R, Harrington H. Specificity of childhood psychotic symptoms for predicting schizophrenia by 38 years of age: a birth cohort study. Psychol Med. 2013;43:2077–86.

50. Zhang W, Zhu Y, Sun M, Guo R, Wu G, Wang Z, et al. Longitudinal trajectories of psychotic-like experiences and their relationship to emergent mental disorders among adolescents: A 3-year cohort study. J Clin Psychiatry. 2019;80:18m12437.

51. Poulton R, Caspi A, Moffitt T, Cannon M, Murray R, Harrington H. Children’s self-reported psychotic symptoms and adult schizophreniform disorder -: A 15-year longitudinal study. Arch Gen PSYCHIATRY. 2000;57:1053–8.

52. Maitra R, Horne CM, O’Daly O, Papanastasiou E, Gaser C, IMAGEN list of authors, et al. Psychotic Like Experiences in healthy adolescents are underpinned by lower fronto-temporal cortical gyrification: A study from the IMAGEN consortium. Schizophr Bull. 2023;49:309–18.

53. Eack SM, Prasad KMR, Montrose DM, Goradia DD, Dworakowski D, Miewald J, et al. An integrated psychobiological predictive model of emergent psychopathology among young relatives at risk for schizophrenia. Prog Neuropsychopharmacol Biol Psychiatry. 2008;32:1873–8.

54. Mustonen A, Niemelä S, Nordström T, Murray GK, Mäki P, Jääskeläinen E, et al. Adolescent cannabis use, baseline prodromal symptoms and the risk of psychosis. Br J Psychiatry. 2018;212:227–33.

55. Aleman A, Kahn RS, Selten J-P. Sex differences in the risk of schizophrenia: evidence from meta-analysis. Arch Gen Psychiatry. 2003;60:565–71.

56. Denissoff A, Mustonen A, Miettunen J, Alakokkare A-E, Veijola J, Scott JG, et al. Trajectories of adolescent psychotic-like experiences and early cannabis exposure: Results from a Finnish Birth Cohort Study. Schizophr Res. 2022;246:95–102.

57. Rimvall MK, van Os J, Verhulst F, Wolf RT, Larsen JT, Clemmensen L, et al. Mental health service use and psychopharmacological treatment following psychotic experiences in preadolescence. Am J Psychiatry. 2020;177:318–26.

