## Supplemental Tables for "Evaluation of Elicit as an adjunct search, screening, and data extraction tool for systematic reviews"

**Supplemental Table 1. Screening Instructions for Elicit**

| Criterion | Input Instructions |
|---|---|
| **Baseline Clinical Status** | Does the study focus on participants without established psychotic disorders at baseline (i.e., subclinical or at-risk populations rather than exclusively diagnosed psychotic disorder patients)? |
| **Baseline Psychosis Spectrum Symptoms** | Does the study assess psychosis spectrum symptoms (subclinical psychotic experiences, attenuated psychotic symptoms, psychotic-like experiences, or clinical high-risk states) at baseline? |
| **Follow-up Duration** | Does the study have at least 12 months of follow-up from baseline assessment? |
| **Population Age** | Do participants have a mean age of 25 years or less at baseline? |
| **Study Size and Design Adequacy** | Is this study design adequate for analysis (i.e., NOT a case report, case series with fewer than 10 participants, or study lacking comparison groups)? |
| **Study Design *** | Is this a prospective longitudinal cohort study, or a case-control study with longitudinal follow-up? (i.e., NOT retrospective studies, controlled clinical trials, cross-sectional studies, case series, dissertations, abstracts, grey literature, and study protocols)? |
| **Diagnostic * Outcomes** | Does the study use validated diagnostic criteria (DSM or ICD), or measures based on DSM or ICD (for example, SCID is based on DSM criteria) to assess mental health disorder outcomes, rather than relying solely on non-validated self-report measures for diagnosis? |
| **Date *** | Was this paper published (publishing date) before August 2025? |
| **Poster *** | The format is NOT a poster. |
| **Systematic Review *** | The format is NOT a systematic review. |

* = Which criteria were set as "strict criterion," which dictates that the answer to the criterion must be "yes," otherwise the paper is automatically excluded.

**Supplemental Table 2. Instructions for Elicit Data Extraction**

| Criterion | Input Instructions |
|---|---|
| **Author** | The authors of the paper |
| **Year** | When was this paper published (the date from their citation) |
| **Average Age** | The average age of the participants of the study at baseline |
| **Community/Help-Seeking** | What was the setting type (community, help-seeking) |
| **Follow-Up Time** | Extract longitudinal study design details for tracking mental health outcomes over time, including:<br>- Total follow-up duration (minimum 1 year as per inclusion criteria), as well as the follow-up duration from the first visit where psychosis spectrum symptoms were assessed to the visit where a diagnosis was made<br>- Number and timing of follow-up assessments<br>- Methods for outcome ascertainment (clinical interviews, registry linkage, self-report)<br>- Loss to follow-up rates and handling of missing data<br>- Whether follow-up was continuous or at discrete time points |
| **Sample Size** | What was the sample size |
| **Female/Male Ratio** | What was the number of males and females in the study |
| **Country** | Where was the study conducted (country) |
| **PSS Assessment Used** | The measure used to assess/gather baseline psychosis spectrum symptoms |
| **Effect Type** | Extract predictive associations, including:<br>- Effect sizes (including Odds Ratios, Hazard Ratios, Risk Ratios)<br>- Statistical significance<br>- Predictive accuracy measures used<br>- Time-dependent effects<br>- Dose-response relationships |
| **Effect** | Extract predictive associations, including:<br>- Effect sizes (including Odds Ratios, Hazard Ratios, Risk Ratios)<br>- Statistical significance<br>- Predictive accuracy measures used<br>- Time-dependent effects<br>- Dose-response relationships |
